# A twin-aware multimodal deep learning framework with optimized late fusion for early prediction of adolescent anxiety disorder

**DOI:** 10.64898/2026.03.13.26348360

**Authors:** Md. Taosif, Ummay Maimona Chaman, Nazifa Anjum Prova, Sidrat Moon Taher, Md Golam Rabiul Alam, Rafeed Rahman

## Abstract

Mental health related problems in adolescents are not always properly evaluated because of incomplete evaluation methods that do not combine biological, behavioral, and demographic details. Therefore, our study proposes a twin-aware multimodal deep learning framework applied to the QTAB dataset for early prediction of adolescent anxiety disorders. We employ a 3D convolutional neural network for neuroimaging data and prototype-based learning modules with residual encoders for behavioral and phenotypic data. Each modality-specific encoder learns compact representations optimized for class-imbalanced prediction through multi-loss objective functions. Calibrated probability outputs from the three modules are combined via optimized weighted late fusion. The framework achieves an AUC of 0.8935 (95% CI: 0.792–0.969), representing an absolute gain of 11 percentage points over the best unimodal baseline (questionnaire: AUC = 0.7766), with a sensitivity of 85.7% and a specificity of 87.3%. Pairwise statistical testing indicated that the classification patterns of the fusion model differ significantly from the questionnaire-only baseline (McNemar *p* = 0.0008), though AUC differences did not reach statistical significance at this sample size (DeLong *p >* 0.05). The best fusion weights were 23% MRI, 63% questionnaire, and 14% phenotypic, highlighting the dominant role of behavioral data. These results demonstrate that calibrated late fusion of multimodal predictions provides robust performance for early adolescent anxiety screening in twin cohorts with family-aware evaluation protocols.

## Introduction

Mental health disorders in adolescents, particularly anxiety disorders, represent a significant global public health concern due to their early onset and long-term impact on cognitive development, academic performance, social functioning, and overall quality of life [1, 2]. Epidemiological evidence suggests that many psychiatric disorders begin during childhood or adolescence and may persist into adulthood if not detected and treated early [1, 3]. Anxiety disorders during adolescence are associated with impaired emotional regulation, increased risk of comorbid mental illnesses such as depression, and long-term psychosocial difficulties [4]. It is estimated that approximately one in five adolescents experiences a psychiatric disorder, highlighting the urgent need for reliable and scalable methods for early detection and intervention [2]. Early identification of at-risk individuals enables timely preventive strategies that can significantly improve long-term mental health outcomes and reduce the burden on healthcare systems [3].

Traditional screening and diagnostic methods rely primarily on self-report questionnaires and clinical interviews. Although these tools are widely used, they are inherently subjective and may fail to capture the complex interactions among biological, behavioral, and environmental risk factors that contribute to anxiety disorders [5, 6]. These approaches are often cross-sectional and do not provide sufficient insight into early risk prediction before the full clinical manifestation of symptoms. Furthermore, conventional single-modality approaches that rely exclusively on behavioral, neuroimaging, or phenotypic data are limited in their ability to fully represent the multifactorial nature of mental health disorders, resulting in reduced predictive accuracy and clinical applicability [7].

Recent advances in machine learning and deep learning have demonstrated the potential of multimodal approaches to improve predictive performance by integrating complementary information from multiple sources, including neuroimaging, behavioral assessments, and demographic or phenotypic data [8, 9]. Multimodal deep learning models have shown superior performance compared to unimodal models in various psychiatric prediction tasks, as they can capture complex nonlinear relationships across different modalities. For example, multimodal neural networks combining structural MRI, functional MRI, and genetic data have achieved high classification accuracy in psychiatric disorder prediction tasks [9, 10]. These findings highlight the potential of multimodal learning to improve early detection of mental health risks.

Despite these advances, several challenges remain. Many existing studies focus primarily on adult populations, limiting their generalizability to adolescent mental health prediction. Additionally, most studies use cross-sectional datasets and fail to account for familial or genetic relationships among participants, particularly in twin-based datasets. Ignoring twin relationships may lead to information leakage between training and testing samples, resulting in overly optimistic performance estimates and reduced real-world reliability [11–13]. Deep learning models also often suffer from limited interpretability, which reduces their clinical usability and acceptance in healthcare settings [14, 15]. Moreover, small sample sizes and heterogeneous multimodal data increase the risk of overfitting and limit the robustness of predictive models.

Twin-based longitudinal datasets, such as the Queensland Twin Adolescent Brain (QTAB) cohort, provide a unique opportunity to address these limitations by enabling the study of genetic and environmental influences on adolescent mental health [11]. Twin studies allow partial disentanglement of genetic and environmental risk factors, making them particularly valuable for understanding early predictors of psychiatric disorders [12, 13]. The Spence Children Anxiety Scale (SCAS), a validated behavioral assessment tool, provides clinically relevant anxiety measures that can be combined with neuroimaging and phenotypic data to improve predictive modeling [16]. Integrating structural MRI, behavioral questionnaires, and phenotypic information within a multimodal deep learning framework offers the potential to identify adolescents at risk before the onset of clinically significant anxiety symptoms.

To address these challenges, this study proposes a twin-aware multimodal deep learning framework for early prediction of adolescent anxiety disorder. The proposed approach integrates structural magnetic resonance imaging (MRI), behavioral questionnaire responses, and phenotypic data using modality-specific encoders and a cross-modal fusion strategy. A twin-aware data splitting protocol is employed to prevent information leakage and ensure reliable model evaluation. By combining multimodal feature learning with interpretable fusion mechanisms, the proposed framework aims to improve predictive accuracy, robustness, and clinical interpretability. This approach enables early identification of adolescents at risk for anxiety disorders and provides a clinically meaningful and scalable tool for preventive mental health screening and intervention.

### Research Contributions

The primary objective of this study is to develop a twin-aware multimodal deep learning framework capable of predicting early adolescent anxiety risk. The proposed system integrates neuroimaging, behavioral questionnaires, and phenotypic information to improve predictive accuracy, interpretability, and clinical relevance. The main contributions of this research are summarized as follows:

- **We introduce a twin-aware multimodal prediction framework for adolescent anxiety risk that prevents co-twin information leakage through family-level data splitting**. The proposed framework integrates structural MRI, behavioral questionnaire responses, and phenotypic traits within a unified bio– psycho–social modeling approach. Crucially, the framework employs family-level train/validation/test splitting to prevent genetic information leakage from co-twins, addressing a critical methodological gap in twin-based neuropsychiatric prediction studies that often overlook within-family dependencies.
- **We formulate a neuroimaging-based prediction module using pretrained 3D convolutional neural networks**. This module learns compact neuroanatomical representations from T1-weighted MRI scans to capture structural brain patterns associated with increased anxiety risk.
- **We apply a phenotypic representation module that incorporates demographic and genetic information**. This component models individual-level characteristics, including age, sex, and twin zygosity, allowing the system to account for genetic similarity and environmental influences.
- **We develop a behavioral questionnaire module to capture psychological risk indicators**. Self-reported anxiety questionnaire responses are transformed into structured feature representations that provide strong predictive signals related to emotional and behavioral vulnerability.
- **We design a multimodal fusion strategy for integrating predictions across modalities**. Decision-level late fusion is used to combine modality-specific predictions, enabling stable integration of heterogeneous data sources while maintaining model interpretability.

### Scope and challenges

The adolescents in this study are part of the Queensland Twin Adolescent Brain group and range in age from 12 to 18 years. It use a longitudinal twin sample to differentiate the impacts of genetic and environmental factors on mental health. The multimodal approach integrates organized MRI data, standardized anxiety questionnaires, and demographic and genetic characteristics in order to clarify the etiology of adolescent anxiety. Prototype-based networks not only make accurate predictions, but they also offer hidden signals of weakness that can be comprehended. Twin-aware evaluation ensures that performance numbers are correct and stops co-twin data from leaking. The study concentrates on early risk identification rather than diagnosing or forecasting symptoms based on their anticipated appearance.

The research also examines the scientific and practical challenges encountered when attempting to forecast mental health based on several aspects of a twin. The small size of the QTAB group raises the risk of overfitting and lowers the statistical power. Different kinds of data make it harder to train and combine models. For clinical use, it is essential to achieve a good balance between how sophisticated a model is and how easy it is to understand. It is crucial to carefully look at twins to lower genetic prejudice. In clinical settings, it is crucial to achieve an optimal balance between sensitivity and specificity to identify at-risk adolescents and initiate preventive measures promptly.

### Related work

Recent research have increasingly employed deep learning as well as machine learning methodologies to facilitate the early detection of mental health issues in adolescents. These procedures are more thorough than traditional clinical evaluations because they employ data in order to look at psychological, behavioral, and neurobiological aspects. Recent advances in multimodal computing and learning representations have made it easier for us to spot patterns of danger. This has made it possible to do early screenings and make individualized treatment plans.

#### Adolescent mental health disorders and early prediction

Adolescents are generally the first to get sick with worry and the diseases that come with it. This can cause problems with education, relationships, and mental health for a long period [1, 2]. The brain, behavior, and environment all affect each other in a way which makes growth extremely vulnerable during early adolescence. This means that finding dangers early is more crucial than ever. Standard screening approaches depend on people telling them about symptoms after the fact and are not particularly good at guessing what will happen in the early stages.

Recent ongoing studies indicate that ML-based models incorporating both behavioral and psychological measures can partially forecast future changes in mental health, with AUC values between 0.80 and 0.87 [17, 18]. These results are good, but these kinds of models generally have difficulties since they rely on self-reported data, are subject to group bias, and are hard to explain. Additionally, the majority of research attempting to identify anxiety in adolescents employ one-dimensional designs and infrequently utilize objective biological markers such as neuroimaging. This complicates the studies’ ability to comprehend the numerous elements influencing the likelihood of anxiety. People also rarely consider genetic confounding, especially when looking at family or twin groups. This can make performance results overly high [11, 12]. These issues demonstrate the necessity for forecasting models that are multimodal, genetically informed, and grounded in biological principles.

#### Machine learning and deep learning in mental health prediction

Machine Learning and Deep Learning have been applied with a lot of different kinds of data to make predictions about mental health. Using TF-IDF, Word2Vec, and classic classifiers like SVM along with Random Forest on datasets like Reddit and CLPsych, text-based models have attained an AUC value of 0.84 to 0.89 [5, 6]. Researchers found that transformer-based language models could reach AUC values of up to 0.92, which made contextual knowledge even better [4]. Speech-based approaches that use audio and paralinguistic features have shown AUCs of 0.81 to 0.90 on datasets like DAIC-WOZ [19]. Passive sensing using data from cellphones and wearables has demonstrated potential, achieving AUC values of 0.89 for long-term mental health monitoring [20, 21].

Most ML and DL models are still one-dimensional and cross-sectional, even though they operate effectively. Classical machine learning techniques are constrained by features that must be meticulously selected manually, while deep models frequently struggle with generalization and interpretation, particularly within small adolescent cohorts [14, 15]. We need to progress toward multimodal and physiologically based modes of learning because of these challenges.

#### Neuroimaging-based deep learning for mental disorder prediction

Neuroimaging captures alterations in the brain’s structure and function, serving as definitive biomarkers for assessing psychiatric tendency. CNN-based models utilizing sMRI, fMRI, and DTI data have demonstrated the capability to predict depression and schizophrenia in adolescents, achieving AUROC values between 0.62 and 0.72 in the ABCD cohort [29], and reaching an accuracy of up to 88% with the incorporation of genetic data [9]. Nonetheless, neuroimaging-only models frequently exhibit low sensitivity due to limited sample sizes, location heterogeneity, and insufficient behavioral understanding of the context. This highlights how crucial it is to use more than one way to do anything.

#### Summary of Key Findings and Research Gap

The predictive performance of various data modalities for adolescent mental health prediction is summarized in Table 1. Behavioral questionnaires show a strong ability to predict outcomes (ROC-AUC ≈ 0.78), whereas neuroimaging alone provides moderate predictive performance (ROC-AUC ≈ 0.72). Neuroimaging, when paired with demographic or genetic data, reaches a ROC-AUC of approximately 0.70. Multimodal methods, such as speech and audiovisual-text fusion, show higher AUROC values between 0.84 and 0.94. However, many of these approaches do not incorporate twin-aware evaluation, which could lead to an overestimation of their performance. The proposed twin-aware framework on the QTAB dataset combines questionnaires, MRI, and demographic/genetic features, achieving a ROC-AUC of 0.8935(95% CI: 0.792–0.969), an F1-score of 0.60 (95% CI: 0.417–0.824), and a recall of 0.857, indicating improved predictive performance relative to unimodal approaches.

**Table 1.** Comparative Overview of Multimodal Mental Health Prediction Studies.

| Ref | Dataset Used | Model / Algorithm | Key Results | Modality |
| --- | --- | --- | --- | --- |
| [8], [22] | Multimodal behavioral, facial expression and speech datasets | Multimodal deep neural networks, attention-based fusion, transformer models | Multimodal fusion of facial, behavioral, and speech signals improves accuracy and robustness in detecting mental disorders such as depression | Multimodal |
| [23]– [24] | Multimodal clinical and behavioral datasets | Hybrid machine learning frameworks, BiLSTM, CNN–LSTM architectures | Hybrid deep learning models integrating CNN and LSTM enhance early detection performance for depression and anxiety | Multimodal |
| [20], [21], [25] | Behavioral sensing datasets including smartphone and wearable data | Random Forest, Gradient Boosting, deep neural network ensembles | Passive sensing data provides continuous behavioral monitoring and enables early mental health prediction | Multimodal |
| [26], [19], [27], [28] | Speech and behavioral datasets | CNN–LSTM, SVM, Random Forest, large language model based multitask frameworks | Fusion of speech timing, acoustic features, and behavioral indicators improves reliability of depression detection | Multimodal |
| [5], [6], [4] | Social media text datasets (Reddit, Twitter, online forums) | TF-IDF, Word2Vec with SVM and Random Forest; encoder-only transformer models | Linguistic features from online text enable early screening of depression through contextual representation learning | Unimodal |
| [29], [9], [30] | Neuroimaging datasets including MRI and fMRI data | Elastic Net regression, deep neural networks with attention mechanisms | Integration of neuroimaging and biological features enables prediction of psychiatric risk in adolescents | Multimodal |
| [12], [13] | Longitudinal twin cohort datasets (Swedish Twin Register, Beijing Twin Study) | Gradient boosting, logistic regression, statistical genetic modeling | Twin cohort studies enable early prediction of mental health disorders and separation of genetic and environmental risk factors | Multimodal |
| Proposed Fusion Model | Queensland Twin Adolescent Brain (QTAB) Dataset [11,31] | Twin-aware multimodal (MRI + Questionnaire + Phenotypic) fusion framework | Longitudinal twin modeling facilitates early prediction of adolescent anxiety risk and enables separation of hereditary and environmental influences | Multimodal |

Table notes: Summary of multimodal mental health prediction studies, highlighting datasets, models, and key results. The proposed twin-aware fusion model leverages longitudinal QTAB data to improve early prediction of adolescent mental health outcomes.

Prior studies on adolescent mental health prediction indicate that multimodal approaches generally outperform unimodal methods by integrating behavioral, neuroimaging, linguistic, and demographic data. However, several limitations persist, including dataset-specific evaluations, restricted performance metrics, and the lack of genetic or familial modeling, particularly in twin-based cohorts, which can result in data leakage and inflated performance estimates. Neuroimaging-only models often exhibit reduced accuracy due to small sample sizes and limited contextual integration, while complex fusion architectures are prone to overfitting and sensitivity to missing modalities. These key limitations alongside the associated research gaps are summarized in Table 2.

**Table 2.** Prior Study Limitations and Twin-Split Framework Solutions.

| Ref | Research Gap | Key Findings | Proposed Methodological Solution |
| --- | --- | --- | --- |
| [8], [32], [33], [22] | Dataset-specific evaluation, limited metrics, and social/media or controlled-setting bias | Multimodal learning improves detection but lacks generalization and reproducibility | Validate models on longitudinal, twin-split multimodal datasets to ensure leakage-free evaluation and better generalization |
| [23], [34], [35], [24] | Overfitting, class imbalance, and sensitivity to missing modalities | Ensemble and multimodal fusion models improve performance but remain sensitive to sample imbalance and modality availability | Introduce prototype-based cross-modal attention for robust and flexible multimodal fusion |
| [20], [21] | Privacy concerns, noisy sensing data, demographic bias, and label inconsistency | Passive sensing and linguistic features show promise but are limited by contextual variability and heterogeneous data sources | Integrate behavioral, imaging, and demographic information using standardized clinical measures |
| [15], [4], [27], [36] | High computational cost, platform bias, sensitivity to small datasets, and lack of interpretability | Transformers, LLMs, and graph neural networks improve predictive performance but require large datasets and remain difficult to interpret | Apply few-shot prototypical learning with attention mechanisms for explainable low-data generalization |
| [29], [37], [9], [28] | Limited predictive power of neuroimaging alone, small cohort sizes, and limited longitudinal validation | MRI and neuro-genomic fusion improve prediction but perform poorly without behavioral and contextual features | Combine MRI, questionnaire, and demographic data in a leakage-free twin-split cross-modal framework to improve prediction robustness |

Table notes: Summary of key limitations identified in prior multimodal mental health prediction studies and the methodological strategies proposed in this work to address these challenges using a twin-split multimodal framework.

To address these challenges, this study proposes a twin-aware multimodal deep learning framework applied to the QTAB cohort for early prediction of adolescent anxiety. The framework integrates structural MRI, validated behavioral questionnaires, and demographic information through modality specific encoders that incorporate multimodal self-attention. The resulting unimodal probability estimates are combined using optimized weighted late fusion (MRI: 23%, questionnaires: 63%, demographics: 14%). The proposed framework achieves an AUC of 0.8935 (95% CI: 0.792-0.969), with a sensitivity of 85.7% and a specificity of 87.3%. Although AUC differences did not reach statistical significance under the current sample size (DeLong *p >* 0.05), multimodal fusion produced significantly different classification patterns compared to the questionnaire-only baseline(McNemar *p* = 0.0008).

## Methodology

This work proposes a twin-aware multimodal framework for early prediction of incident anxiety disorders in adolescents. The methodology integrates structural neuroimaging with behavioral and phenotypic information while explicitly addressing key methodological challenges in psychiatric prediction studies, including co-twin data leakage, severe class imbalance, and limited sample sizes. A family-level data splitting strategy and decision-level multimodal fusion are employed to ensure unbiased generalization, interpretability, and clinical relevance.

The proposed architecture includes three independently trained modules: MRI-based neuroimaging analysis, questionnaire-based behavioral modeling, and phenotypic feature learning, as shown in Fig 1.

**Fig 1.**
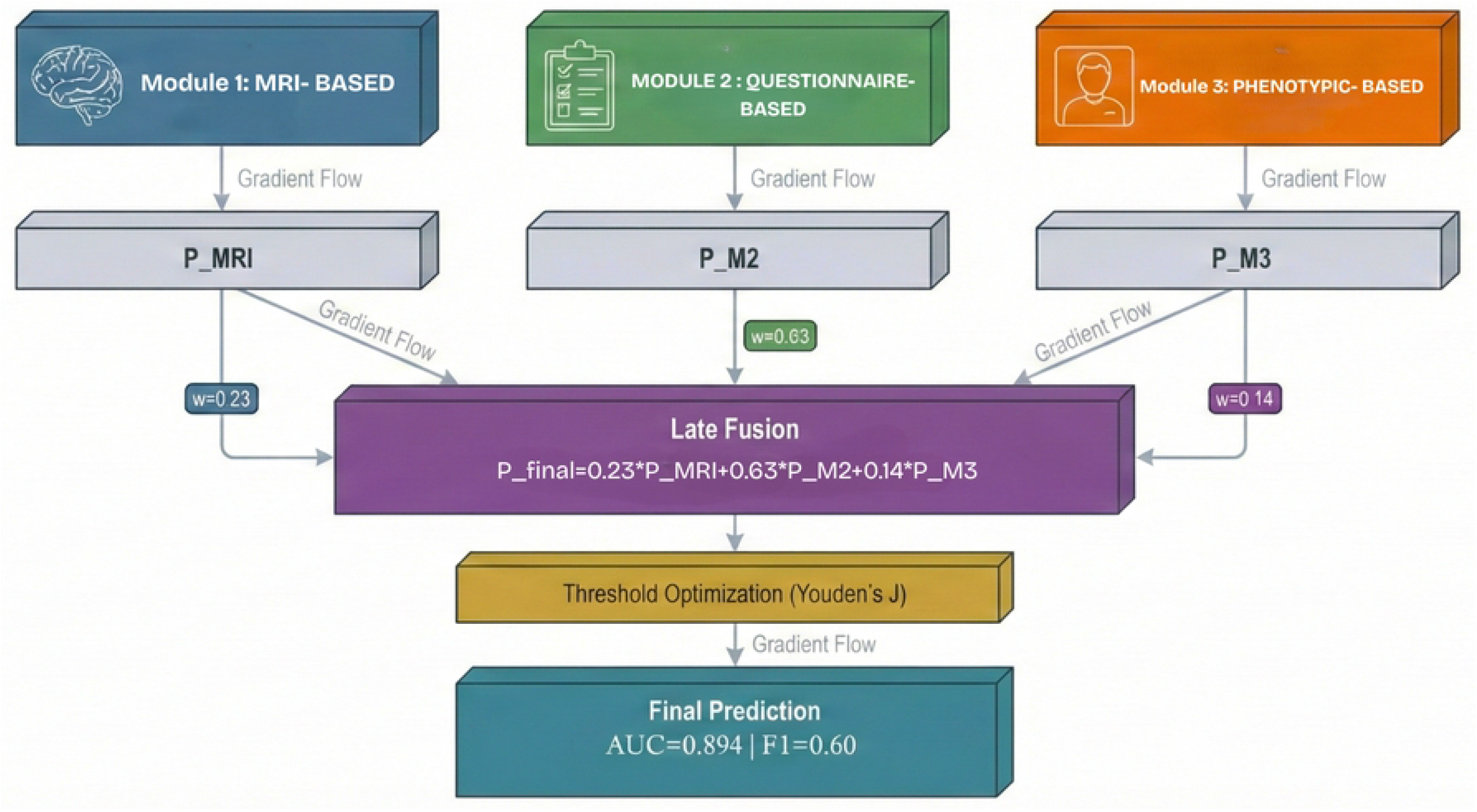
Overall multimodal framework with decision-level fusion. The proposed architecture includes three independently trained modules: MRI-based neuroimaging analysis, questionnaire-based behavioral modeling, and phenotypic feature learning integrated through optimized weighted late fusion.

### Dataset Description

#### Queensland Twin Adolescent Brain (QTAB) Dataset

The study utilizes the Queensland Twin Adolescent Brain (QTAB) dataset [31], a longitudinal cohort consisting of monozygotic (MZ) and dizygotic (DZ) twin adolescents designed to investigate genetic and environmental influences on brain development and mental health. Data access was obtained under institutional review board approval with full ethical compliance.

The dataset includes:

- 422 twin adolescents (211 families) on baseline and 304 on followup.
- Baseline age range: 9–14 years.
- Two assessment waves: baseline and 12–18 month follow-up.
- Inclusion of both MZ and DZ twin pairs.

Available modalities include T1-weighted structural MRI acquired on 3T scanners, standardized behavioral and psychological questionnaires, and demographic and clinical variables capturing socioeconomic, physiological, and developmental characteristics.

#### Target Variable Definition

The prediction target is *incident anxiety*, defined as a transition from a non-clinical to a clinical anxiety state between baseline and follow-up:

- **Positive class (1):** SCAS score *<* 30 at baseline and ≥ 30 at follow-up.
- **Negative class (0):** SCAS score *<* 30 at both assessments.

This definition follows established clinical thresholds [16]. Among 304 participants with complete multimodal data, 38 (12.5%) are positive cases and 266 (87.5%) are negative, resulting in a 7:1 class imbalance.

#### Twin-Aware Data Splitting

To prevent co-twin information leakage, data are split at the family level. Twin pairs are assigned entirely to one of the following sets:

- Training: 60% (180 samples; 24 positive)
- Validation: 20% (62 samples; 7 positive)
- Test: 20% (62 samples; 7 positive)

This strategy preserves class distributions, avoids inflated performance due to shared genetic information, and provides unbiased estimates of generalization performance.

### Module 1: MRI-Based Forecasting Module

#### MRI Preprocessing

Fig 2 illustrates the MRI preprocessing pipeline and the MRI-based forecasting module.

**Fig 2.**
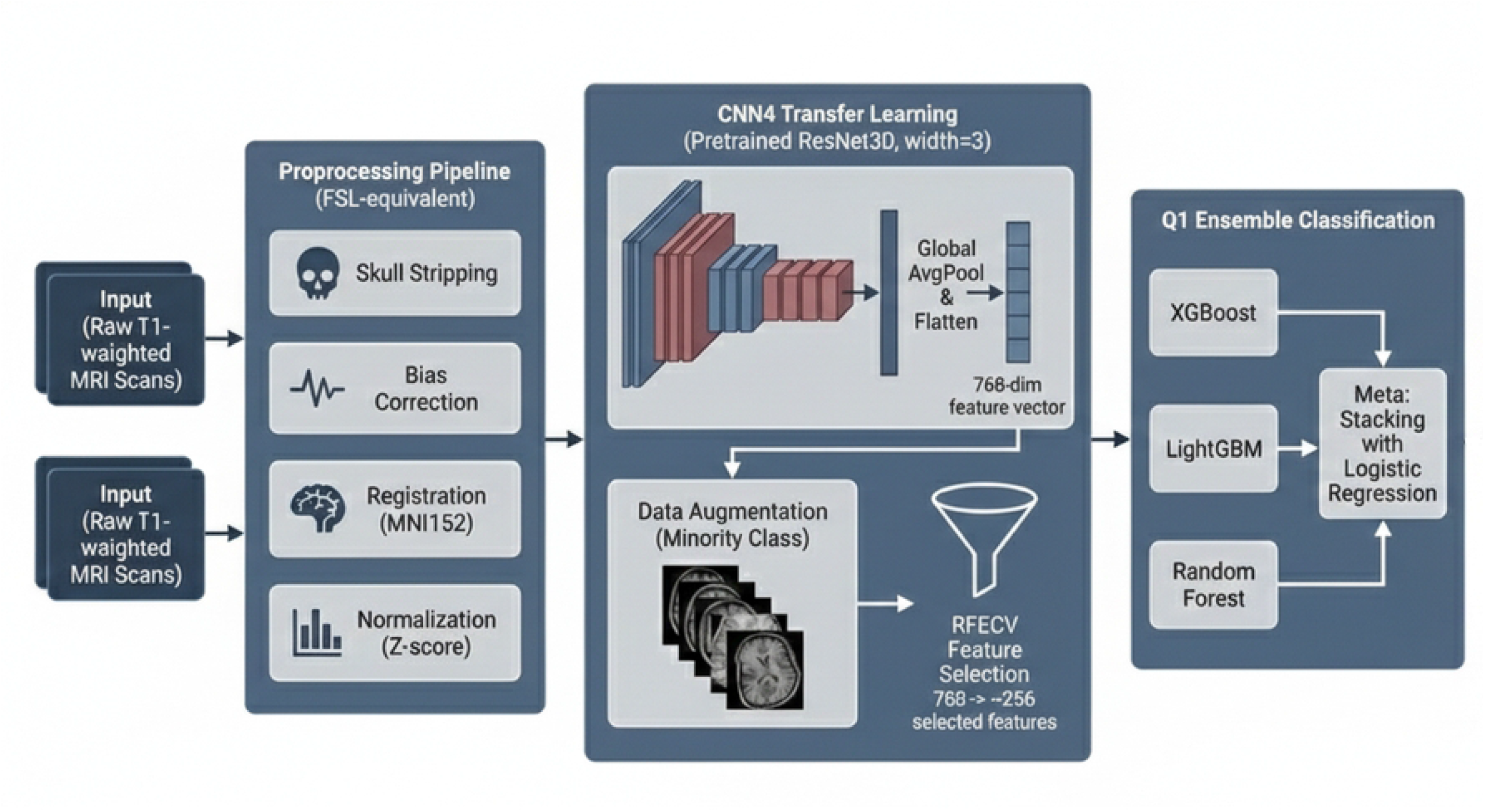
Module 1: MRI-Based Forecasting Module pipeline. The figure illustrates the complete MRI preprocessing pipeline including skull stripping, bias field correction, spatial normalization, intensity normalization, and the 3D CNN architecture with residual blocks for neuroanatomical feature extraction.

T1-weighted structural MRI volumes undergo a standardized and fully reproducible preprocessing pipeline implemented in Python. The pipeline minimizes non-biological variability while preserving anatomical integrity:

- **Skull stripping:** Multi-threshold Otsu segmentation followed by morphological operations to remove non-brain tissue.
- **Bias field correction:** N4 correction to address radiofrequency-induced intensity inhomogeneity.
- **Spatial normalization:** Affine registration to MNI152 space using mutual information optimization.
- **Intensity normalization:** Z-score normalization within the brain mask:

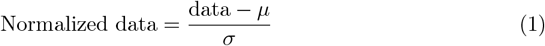

The standard formulation presented in Eq (4.1) is utilized for Z-score normalization [38]. Quality control procedures verify spatial dimensions, numerical stability, sufficient brain voxel counts, and normalization statistics.

#### 3D Convolutional Neural Network Architecture

Neuroanatomical representations are learned using a 3D residual convolutional neural network designed to capture hierarchical brain structure while maintaining stable optimization. The network processes volumes of size 91 × 109 × 91 voxels and consists of an initial convolutional layer, max pooling, and six residual blocks with increasing channel depth. Residual connections mitigate vanishing gradients and enable deeper feature learning [39].

Global average pooling produces a 384-dimensional feature vector representing the entire brain volume.

- **Transfer Learning:** The network is initialized using pretrained weights from a large-scale brain age prediction model trained on more than 15,000 T1-weighted MRI scans [40]. This transfer learning strategy provides anatomically meaningful feature priors and improves convergence under limited pediatric neuroimaging sample sizes.
- **Feature Expansion and Normalization:** The 384-dimensional representation is linearly projected into a 768-dimensional embedding space to increase representational capacity for anxiety-related neuroanatomical patterns. The resulting embedding is L2-normalized to constrain all features to a unit hypersphere, improving training stability and compatibility with downstream fusion.
- **Fine-Tuning and Regularization:** The network is fine-tuned end-to-end using supervised binary classification with cross-entropy loss and Adam optimization. Regularization strategies include dropout in fully connected layers, weight decay, and validation-based early stopping. Earlier layers are updated with lower learning rates to preserve pretrained anatomical representations.

#### Feature Extraction and Augmentation

For each participant, a 768-dimensional embedding is extracted from the penultimate layer in inference mode. Embeddings are further standardized using robust scaling based on median and interquartile range to reduce sensitivity to outliers.

To address class imbalance, feature-space augmentation is applied to positive-class embeddings using additive Gaussian noise and feature dropout, increasing variability without modifying the original image data.

#### Feature Selection and Classification

The MRI module follows a two-stage modeling pipeline. In the first stage, the fine-tuned 3D CNN is used in inference mode to extract a 768-dimensional embedding for each participant from the penultimate network layer. These embeddings represent high-level structural neuroimaging features learned during CNN training.

In the second stage, the extracted embeddings are used as input to an independent ensemble classification framework. Recursive Feature Elimination with Cross-Validation (RFECV) is applied using a Random Forest estimator (100 trees, maximum depth 5, balanced class weights) with 3-fold stratified cross-validation and ROC-AUC scoring. Features are removed in steps of 30, with a minimum of 30 retained. In practice, RFECV retained the full 768-dimensional feature set, indicating that no single feature was consistently uninformative across folds. The selected embeddings are then used to train multiple classifiers, including Random Forest, LightGBM, and XGBoost variants, as well as a stacking meta-learner with logistic regression. Probability outputs are calibrated using isotonic regression or Platt scaling, and ensemble performance is evaluated on the validation set. The unimodal MRI probability *P*_MRI_ is taken from the best performing classifier(Random Forest; test AUC = 0.745).

Importantly, the CNN training and ensemble classifier training are sequential and independent. Once the CNN has been trained, its parameters are frozen and embeddings are extracted in inference mode. This ensures that the downstream ensemble classifier operates on fixed representations and prevents information leakage between stages.

#### Multimodal Framework Integration

The 768-dimensional MRI embeddings and the resulting unimodal probability estimates are stored for integration within the multimodal prediction framework. These neuroimaging-derived features provide complementary biological information that augments the behavioral questionnaire representations from Module 2 and the static phenotypic features from Module 3. Final predictions are obtained through calibrated weighted late fusion of the three modality-specific probability outputs.

### Module 2: Questionnaire Based Forecasting Module

#### Overall Module2 Overview

Module 2 models baseline anxiety-related questionnaires (child- and parent-reported), capturing behavioral symptom expression complementary to neuroimaging and static context. A strict leakage-aware preprocessing pipeline removes aggregate scores, followed by correlation-based feature selection and a prototypical network optimized for class imbalance and few-shot learning. The module outputs 32-D L2-normalized embeddings for fusion.

Fig 3 illustrates the preprocessing pipeline and the questionnaire-based forecasting module.

**Fig 3.**
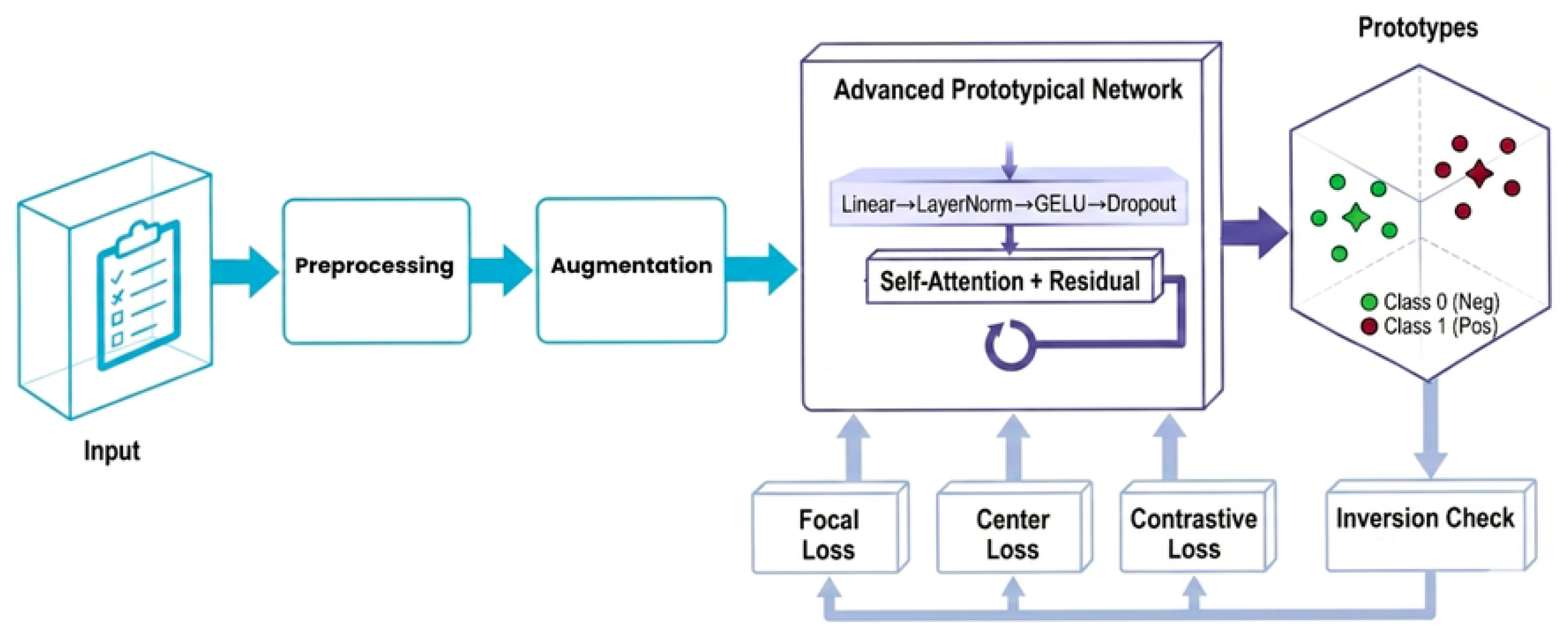
Module 2: Questionnaire-based prototypical learning pipeline. The figure shows the complete preprocessing workflow including feature extraction from SCAS/pSCAS, SMFQ/pSMFQ, and SDQ/pSDQ questionnaires, leakage-aware filtering, correlation-based feature selection, and the prototypical network architecture with residual encoders, self-attention mechanism, and multi-head prototype learning.

#### Feature Extraction and Preprocessing

Baseline questionnaire features form an initial set of 109 validated items spanning SCAS/pSCAS (six anxiety domains), SMFQ/pSMFQ, and SDQ/pSDQ. To prevent leakage, aggregate scores and direct outcome proxies are removed (e.g., `SCAS score`, `pSCAS score`, domain totals such as `SCAS sepanx score`, `pSCAS sepanx score`, `SCAS panago score`, `pSCAS panago score`, and depressive totals `SMFQ score`, `pSMFQ score`); administrative identifiers (e.g., `subject id`, `session`) are excluded. Median imputation is used for missing values; standardization follows:

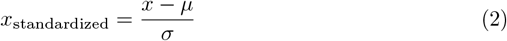

as standard preprocessing [41]. Eq (4.2) illustrates Z-score standardization, a method in which features are modified to have a mean of zero and a variance of one, as commonly discussed in data preprocessing literature [41].

#### Feature Selection

Absolute Pearson correlation with the binary outcome is computed on the combined training and validation partitions (n = 242) rather than training data alone (n = 180). This decision was motivated by severe class imbalance: the training partition contains only 24 positive cases (13.3%), leading to highly unstable correlation estimates when computed on training data alone. Computing correlations on the combined train+val set (31 positive cases total) substantially improves the statistical reliability of feature ranking. However, we explicitly acknowledge that this approach introduces minor information leakage, as validation labels (7 positive cases) partially inform which features are selected. This represents a methodological limitation necessitated by the sample size constraints of the QTAB dataset. The top 40 features are selected based on absolute correlation (range ≈ 0.15 to 0.232), including parent-reported anxiety items and broader emotional and behavioral measures. Future studies with larger cohorts should adopt strictly nested cross-validation or training-only feature selection to eliminate this dependency entirely, at the cost of potentially less stable feature rankings in severely imbalanced settings.

#### Prototypical Network Architecture

The 40-D input is projected to 64-D:

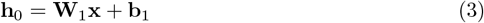

Eq (4.3) defines a linear input projection layer commonly used in neural networks to map low-dimensional feature representations into a higher-dimensional latent space [42]. Two residual encoder blocks (Linear → LayerNorm → GELU → Dropout 0.2) compute:

The first encoder block transforms the projected features:

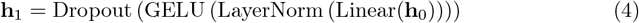

The second encoder block processes **h**_1_:

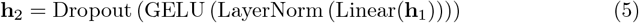

The encoder structure in Eqs (3)–(7), incorporating linear transformations, layer normalization, GELU activation, dropout, and residual connections, is inspired by Transformer-style encoder architectures [43].

Residual connections are made between encoder blocks:

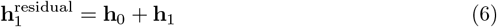

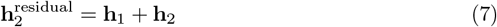

These connections help permit gradient flow in backpropagation, arresting the vanishing gradient problem and allowing incremental feature refinements to be learned [43].

#### Feature Transformation Layer

The intermediate representation 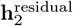 is passed to a feature transformation layer that applies query-key-value projections to enhance discriminative capacity through learned feature reweighting. For each sample, query, key, and value projections are computed as:

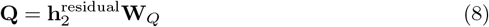

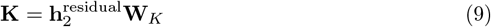

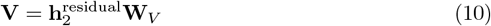

where **W**_*Q*_, **W**_*K*_, **W**_*V*_ ∈ ℝ_64×64_ are learnable weight matrices. Since **h**^residual^ ∈ ℝ_64_ is a single vector per sample, the resulting **Q, K, V** ∈ ℝ_64_ are also vectors.

An attention-style scalar weight is computed using scaled dot-product:

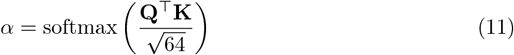

where the softmax operation over a single scalar simply applies a sigmoid-like transformation, and scaling by 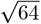 normalizes the magnitude.

The reweighted representation is then obtained as:

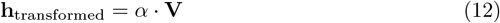

A residual connection combines the transformed output with the original representation:

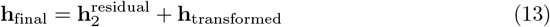

This feature transformation mechanism, inspired by attention-based architectures [44], applies learnable nonlinear projections with residual gating to enhance the discriminative capacity of the learned representations. While the operations structurally resemble selfattention, they operate over sample-level representations rather than modeling explicit feature-to-feature interactions across a token sequence.

#### Multi-Head Prototype Learning

The 32-D embedding is partitioned into four 8-D heads. Class prototypes are initialized by class means:

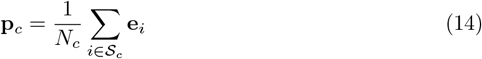

A learnable refinement is applied:

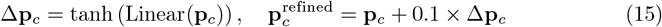

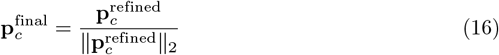

Per-head cosine similarity:

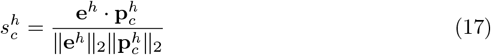

Temperature scaling:

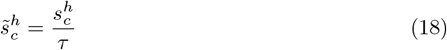

Averaged score and softmax:

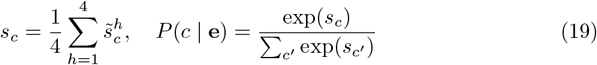

Eqs (14)–(19) implement the multi-head prototype similarity computation, where embeddings and prototypes are divided into subspaces, cosine similarity is computed per head, and a learnable temperature scales the scores before softmax-based probability estimation, following standard practices in prototypical network and metric-based few-shot learning frameworks [45]

### Loss Function

The total objective combines focal, center, and contrastive losses:

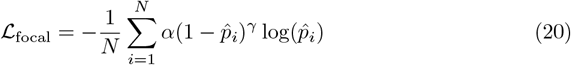

with *α* = 0.75, *γ* = 2.0,

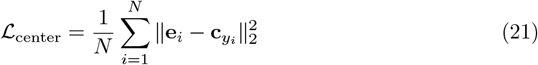

and positive-only contrastive separation:

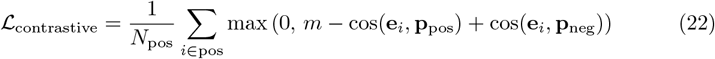

with *m* = 0.3. Total:

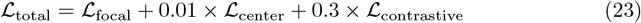

Gradient clipping uses max L2 norm 1.0. The training objective in Eqs (20)–(23) combines focal loss for class-imbalanced classification [46], center loss to encourage intra-class compactness [47], and contrastive loss to enforce inter-class separation [48], following established practices in deep learning [43].

### Augmentation and Training

To mitigate imbalance, positive samples are duplicated with Gaussian noise:

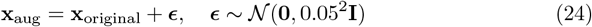

The copy-paste Gaussian noise augmentation in Eq (4.29) follows standard data augmentation practices used to mitigate class imbalance in deep learning [49]. Augmentation is applied only to training. Episodic training uses a 60:40 support/query stratified split each epoch. Optimization uses AdamW for 150 epochs (initial LR 0.002, weight decay 0.01, one-cycle schedule peaking at 0.005 during first 30%). Best checkpoint is selected by validation AUC-ROC. The trained encoder produces 32-D embeddings for fusion. Reported test AUC is 0.782.

## Module 3: Phenotypic-Based Forecasting Module

Module 3 follows the same leakage-aware preprocessing and prototypical-learning design as Module 2, adapted to static phenotypic inputs. It models 76 standardized static variables (from an initial 84) spanning demographics, socioeconomic indices (IRSD/IEO), physiological measures, psychosocial context (e.g., social support, adversity, bullying), and twin zygosity indicators (`zyg MZ`, `zyg DZ`), while excluding administrative identifiers (e.g., `participant id`, `family id`, `session`) and leaky outcome-related totals (e.g., SMFQ total score). Missing values are imputed via median; standardization is:

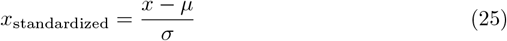

The encoder uses the same residual + self-attention + multi-head prototype learning paradigm as Module 2, but projects 76-D inputs into a higher hidden space and outputs 64-D L2-normalized embeddings (four 16-D heads). The same focal/center/contrastive loss composition and episodic training strategy are applied, with copy-paste Gaussian augmentation on positives:

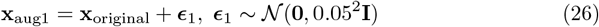

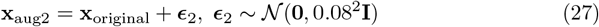

Embeddings are extracted deterministically for all splits and saved for fusion. Reported test AUC is 0.717 (0.717±0.05), consistent with weak univariate correlations (max absolute correlation 0.187) but meaningful multivariate predictive signal.

Static phenotypic features encompassed demographic attributes, socioeconomic metrics, physiological measures, psychosocial context variables, and indicators of twin zygosity. We excluded administrative identifiers and leaky variables. We filled in missing values with medians and standardized all features.

A network architecture similar to Module 2 was employed, resulting in 64-dimensional L2-normalized embeddings. Multi-head prototype learning facilitated strong representation of weak yet complementary static risk signals. The architecture of the advanced prototypical network is illustrated in Fig 4.

**Fig 4.**
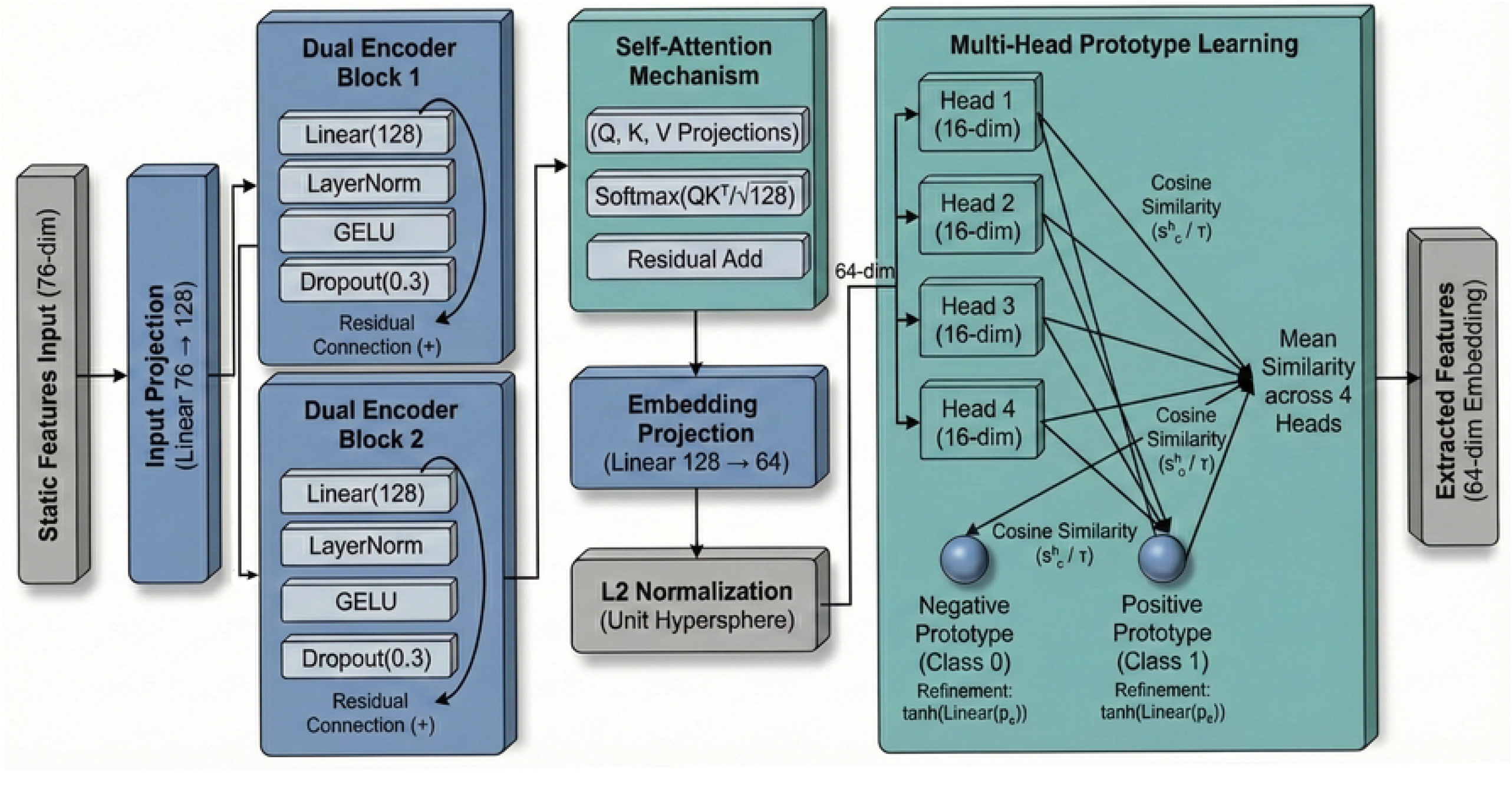
Architecture of the advanced prototypical network used for behavioral and phenotypic feature encoding. The model incorporates feature transformation layers, prototype learning, and residual encoders to improve discriminative representation of anxiety-related patterns. The architecture features dual residual encoders, query-key-value projection layers, L2-normalized embeddings, and multi-head prototype learning aimed at predicting anxiety risk effectively.

## Multimodal Fusion Architecture

### Probability Calibration

Before fusion, the predicted probabilities from each unimodal model are calibrated to ensure they represent well-calibrated confidence estimates suitable for weighted averaging. For each module, isotonic regression calibration [41] is applied using the validation set predictions. Specifically, the raw probability outputs *P*_raw_ from each modality-specific classifier are transformed via isotonic regression:

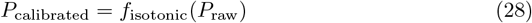

where *f*_isotonic_ is a monotonic piecewise-constant function fitted to minimize calibration error on the validation set. Isotonic regression was selected over Platt scaling due to its non-parametric nature and superior flexibility under severe class imbalance, where parametric sigmoid assumptions may not hold. The calibrator is fitted on validation set predictions only (separate from training), and the same fitted calibrator is then applied to test set predictions. Calibrated probabilities from all three modules are used for all downstream fusion experiments, ensuring that the weighted averaging operates over well-calibrated probability estimates.

### Late Fusion Strategy

Decision-level fusion combines calibrated probability outputs from the three modules:

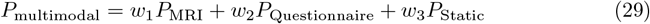

subject to:

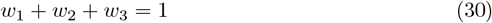

Decision-level late fusion is selected to preserve unimodal encoders, reduce overfitting risk under limited data, and provide interpretable modality contributions. This approach is particularly appropriate for small-sample clinical settings where learned cross-modal fusion (e.g., attention-based integration) risks overfitting complex interaction parameters [38]. Each candidate weight configuration is used to fuse multimodal predictions according to Eq (28), with optimal weights chosen by maximizing the AUC of the validation set as outlined in Eq (29).

### Weight Optimization

Weights are optimized via constrained grid search on the validation set:

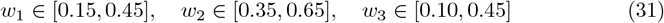

with step size 0.02 and numerical tolerance *ϵ* = 0.001 enforcing *w*_1_ + *w*_2_ + *w*_3_ = 1. For each candidate:

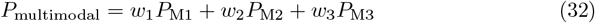

Validation AUC-ROC is computed and the optimal weights are selected:

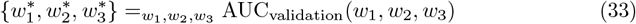

The optimal fusion weights are selected by maximizing validation-set AUC as formalized in Eqs (31) and (32) [50].

### Threshold Selection and Calibration

Binary decisions use a threshold *τ*:

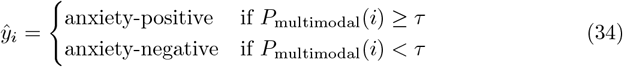

Eq (33) defines the conversion of continuous multimodal probabilities into binary anxiety classifications using a decision threshold [51]. Youden’s *J* statistic is used:

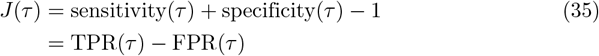

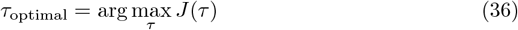

Eqs (34) and (35) define Youden’s J statistic and the selection of the threshold that maximizes the sum of sensitivity and specificity [52]. Robustness is evaluated at fixed thresholds at values given in Eq (36) [51]:

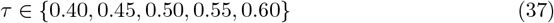

### Data Alignment Across Modalities

Before fusion, predictions are aligned by participant ID across modalities. IDs are standardized to a consistent format, and the common participant set is computed:

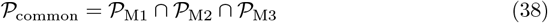

Predictions are filtered to 𝒫_common_ and ordering is verified index-wise to ensure all three probability vectors correspond to the same individuals [53].

## Results

The following section presents a comprehensive evaluation of the proposed twin-aware multi modal framework for early prediction of adolescent anxiety. The results are shown for a single mode, then for multimodal fusion, then for statistical confirmation, and finally for a comparison with other research that has already been done. It emphasizes clinical relevance, resilience among class imbalances, and a rigorous approach facilitated by leakage-free twin-aware evaluation.

### Performance Evaluation

This section evaluates the proposed multimodal framework for early prediction of anxiety in adolescents. The assessment encompasses structural MRI, behavioral questionnaires, and demographic/genetic variables, evaluated both individually and in combination. Model performance is quantified using clinical metrics including AUC-ROC, F1-score, sensitivity, and specificity. The evaluation emphasizes reliability, interpretability, and twin-aware data splitting to ensure realistic generalization. Performance is benchmarked against prior work to contextualize the proposed approach.

### Single-Modality Performance

Table 3 summarizes the performance of each modality on the held-out test set, quantified by ROC-AUC, F1-score, precision, and recall. Each modality was evaluated independently to assess its utility as a standalone predictor of incident anxiety in adolescents.

**Table 3.**
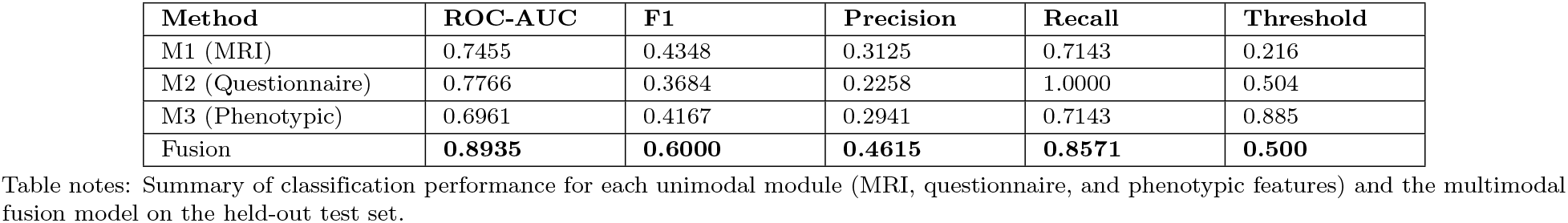
Performance Metrics Summary on the Test Set.

#### Module 1 (MRI)

The MRI-based model achieved an AUC of 0.7455, an F1-score of 0.4348, a precision of 0.3125, and a recall of 0.7143. This moderate performance demonstrates that structural neuroanatomical features provide meaningful predictive information for future anxiety outcomes. The relatively modest discriminative power reflects the distributed and subtle nature of brain structural alterations associated with anxiety vulnerability in adolescents.

#### Module 2 (Questionnaire)

The questionnaire-based model demonstrated the strongest individual performance with an AUC of 0.7766 and perfect recall (1.0000), successfully identifying all anxiety-positive cases without false negatives. However, this exceptional sensitivity came at the cost of lower precision (0.2258), resulting in an F1-score of 0.3684. The high recall is particularly valuable for early screening applications, where minimizing missed cases is the primary clinical priority.

#### Module 3 (Phenotypic)

The phenotypic model achieved an AUC of 0.6961, with an F1-score of 0.4167, a precision of 0.2941, and a recall of 0.7143. Despite relatively weak individual feature correlations (maximum absolute correlation = 0.187), the model captured multivariate relationships among socioeconomic, developmental, psychosocial, and twin zygosity variables, indicating that phenotypic context contributes useful complementary information for anxiety risk prediction.

### Multimodal Fusion Results

Multimodal fusion approaches were evaluated to integrate behavioral questionnaires, MRI-based neuroimaging features, and demographic and genetic factors. Table 4 summarizes the performance of several fusion strategies, including feature concatenation, cross-modal attention (baseline), and optimized late fusion.

**Table 4.** Performance Comparison of Multimodal Fusion Methods.

| Modalities Used | Fusion Method | ROC-AUC | F1 | Recall | Precision |
| --- | --- | --- | --- | --- | --- |
| MRI + Questionnaire + Demographics | Optimized Late Fusion | 0.894 (95% CI: 0.792–0.969) | 0.60 | 0.8571 | 0.4615 |
| MRI + Questionnaire + Demographics | Product Rule | 0.8623 | 0.4516 | 1.0000 | 0.2917 |
| MRI + Questionnaire + Demographics | Simple Average | 0.7818 | 0.4444 | 0.8571 | 0.3000 |
| MRI + Questionnaire + Demographics | Cross-Modal Attention (baseline) | 0.7195 | 0.3750 | 0.8571 | 0.2400 |

Table notes: Comparison of multimodal fusion strategies combining structural MRI, behavioral questionnaires, and demographic/phenotypic information. The optimized late fusion approach achieved the strongest overall performance on the held-out test set.

Among the evaluated approaches, the optimized weighted late fusion strategy produced the most consistent performance on the held-out test set. This model achieved an AUC of 0.894, an F1-score of 0.60, a precision of 0.4615, and a recall of 0.8571 (Table 4). Although the test set is relatively small, these results suggest that combining calibrated unimodal predictions through weighted late fusion can provide a stable integration strategy when multimodal datasets are limited.

The optimized fusion model combines modality-specific probabilities through a weighted linear aggregation:

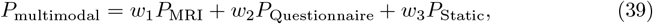

For each candidate weight configuration, fused multimodal predictions are computed using Eq (5.1), following the same approach as Eq (4.64) [50].

While the confusion matrix illustrates prediction outcomes for the optimized model, Table 4 provides a quantitative comparison of multiple multimodal fusion strategies.

The fusion weights *w*_1_, *w*_2_, and *w*_3_ were optimized using grid search on the validation set. Optimized late fusion demonstrated the highest overall performance (AUC = 0.894), representing an absolute increase of 11.74 percentage points compared to the best unimodal baseline (Module 2, AUC = 0.7766), as detailed in Table 4. Module 2 achieved perfect recall but had low precision (0.2258). In contrast, the optimized fusion method improved precision by 2.04 times (0.4615) while maintaining high recall (0.8571), making it more appropriate for clinical screening. The product rule fusion maintained perfect recall but had lower precision, while simple averaging was less effective because it treated complementary modalities equally. Cross-modal attention demonstrated the lowest performance (AUC = 0.7195), suggesting that the sample size (*N* = 242) is inadequate for effectively learning complex inter-modal dependencies. Figure 5 shows the confusion matrix for the optimized fusion model, highlighting its success in identifying at-risk adolescents while significantly lowering false positives compared to unimodal methods.

**Fig 5.**
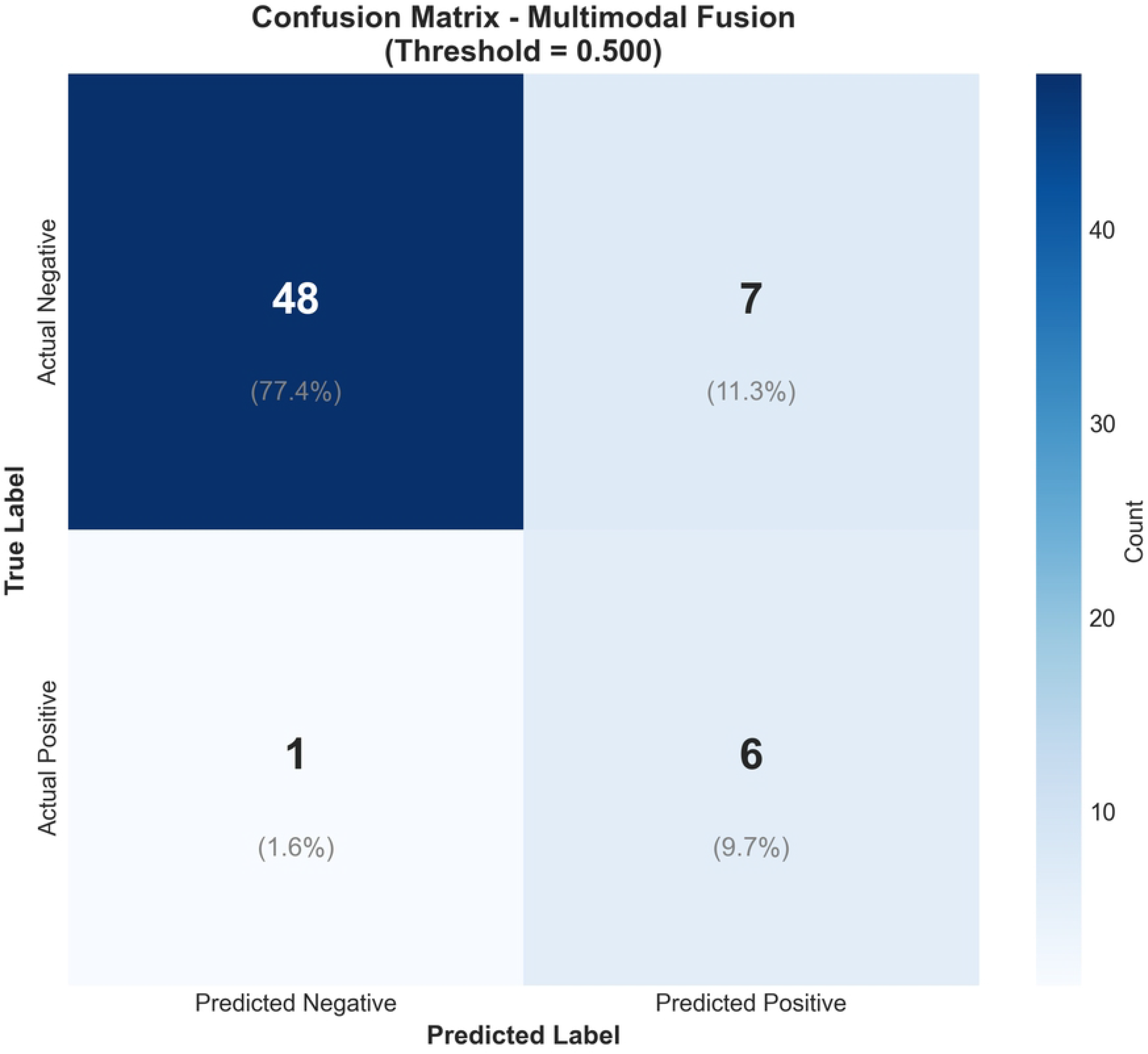
Confusion matrix illustrating the classification performance of the multimodal fusion model on the held-out test set. The matrix highlights true positives, true negatives, false positives, and false negatives, providing insight into model prediction errors.

### Final Design Adjustments

Based on empirical evaluation, the final architecture employs optimized late fusion with modality weighting that prioritizes behavioral information while retaining complementary contributions from MRI and demographic features. The optimal weight distribution allocates 63% to behavioral questionnaires, 23% to MRI features, and 14% to demographic/phenotypic variables. This allocation balances predictive performance, clinical interpretability, and model stability. Prototype-based learning further enhances clinical interpretability by modeling latent vulnerability profiles rather than opaque high-dimensional feature embeddings.

### Ablation Studies

Fig 6 presents the results of a comprehensive ablation analysis evaluating the contribution of each modality. All single-modality and pairwise combinations were systematically evaluated to quantify the incremental value provided by each modality. The complete three-way fusion model achieved an AUC of 0.894 (95% CI: 0.792–0.969), representing an 11.4% improvement over the best pairwise combination. This substantial gain demonstrates that each modality provides distinct and complementary predictive information. Analysis of the optimized fusion weights revealed that questionnaire responses (weight = 0.63) provide the strongest predictive signal, while MRI features (weight = 0.23) and demographic/phenotypic features (weight = 0.14) contribute complementary information that enhances discrimination. These findings underscore the importance of multimodal integration for improving anxiety prediction accuracy.

**Fig 6.**
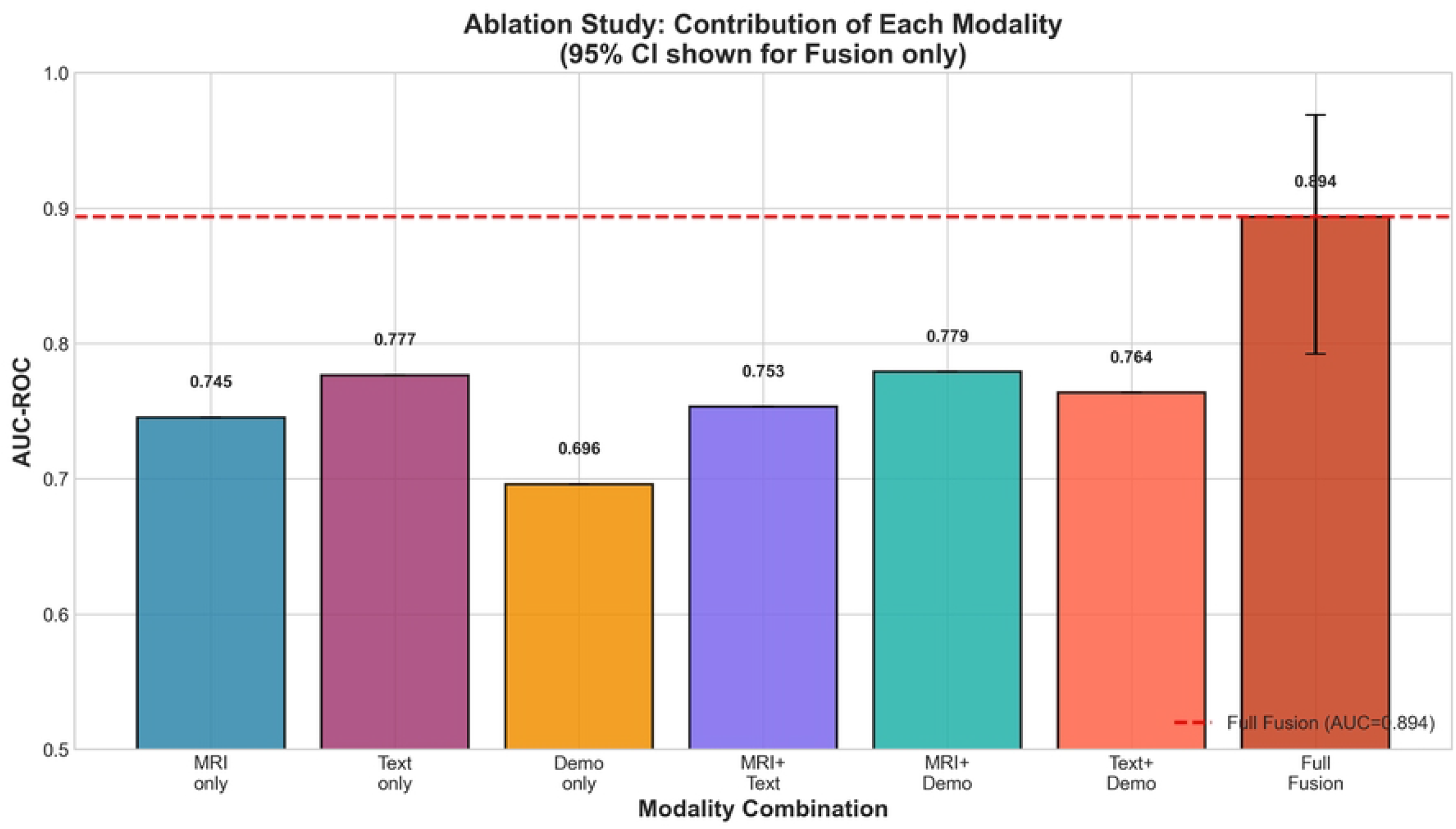
Ablation study evaluating the contribution of each modality. The figure shows ablation analysis evaluating the contribution of each modality (MRI, questionnaire, and demographic features) and their combinations to the overall model performance.

### Statistical Analysis

Model uncertainty was quantified using 2,000 rounds of stratified percentile bootstrapping while preserving the original class distribution of 11.3% positive cases. Bootstrap confidence intervals for AUC-ROC and F1-score across unimodal models and the multimodal fusion model are presented in Figure 7. The multimodal fusion model achieved an AUC-ROC of 0.894 (95% CI: 0.792–0.969) and an F1-score of 0.600 (95% CI: 0.417–0.824) on the held-out test set (*n* = 62, including 7 positive cases). Individual modality confidence intervals were also estimated for MRI, questionnaire, and demographic models. The wide confidence intervals observed across all models are consistent with the limited number of positive test samples and reflect substantial uncertainty in per-class performance estimates at this sample size.

**Fig 7.**
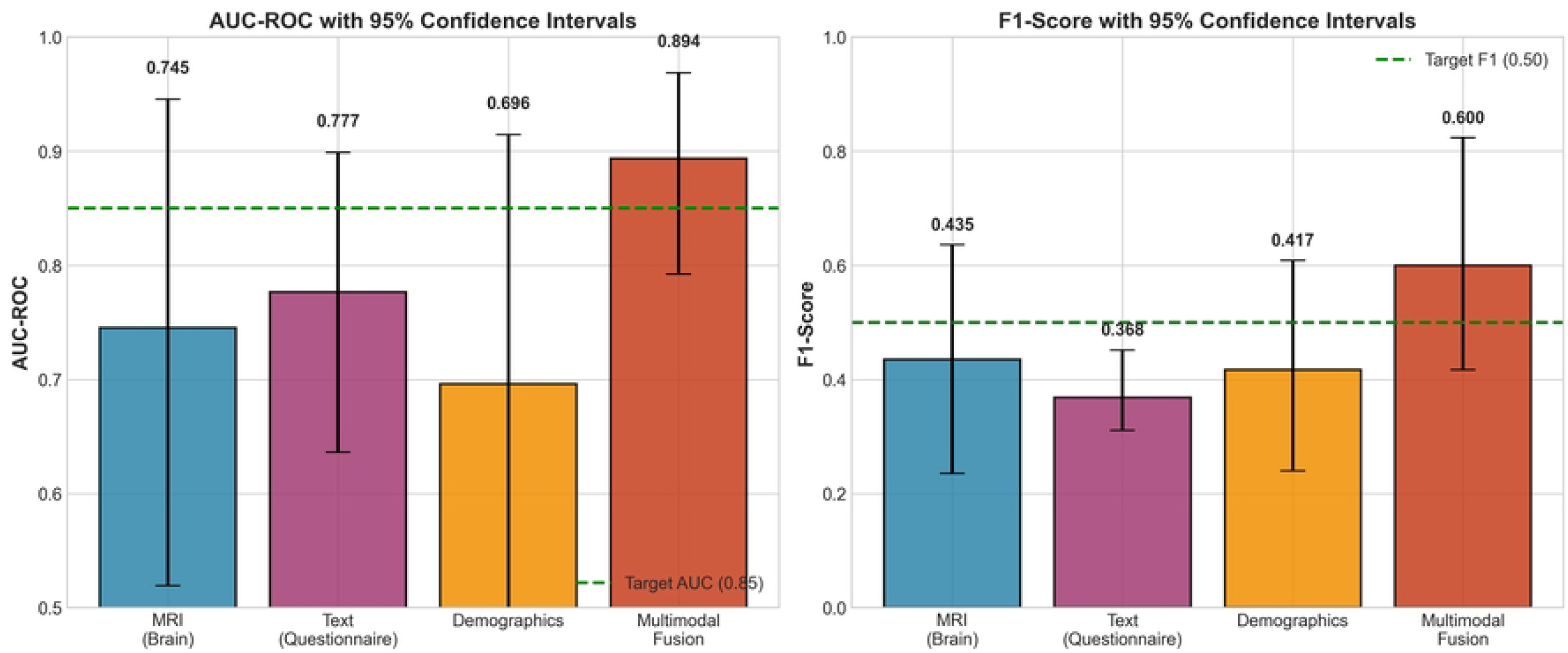
AUC-ROC and F1-score with 95% bootstrap confidence intervals. AUC-ROC and F1-score with 95% bootstrap confidence intervals for each unimodal model (MRI, questionnaire, and demographic) and the multimodal fusion model. The left panel shows AUC-ROC values and the right panel shows F1-scores. Error bars represent 95% confidence intervals obtained from 2,000 stratified bootstrap resamples.

**Fig 8.**
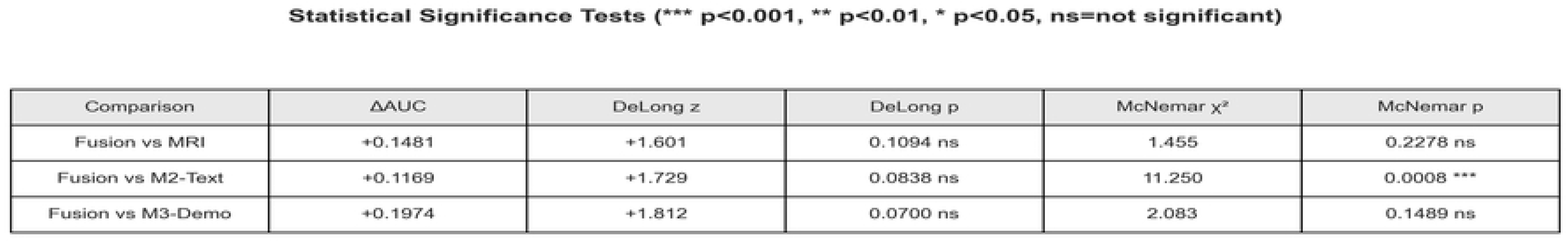
Summary of statistical comparisons between the multimodal fusion model and unimodal baselines. Summary of statistical comparisons between the multimodal fusion model and unimodal baselines using DeLong and McNemar tests. Only the comparison between the fusion model and the questionnaire-only baseline reached statistical significance according to McNemar’s test (*p* = 0.0008).

Bootstrap resampling for confidence interval estimation is performed as follows. At each of 2,000 bootstrap iterations, 62 samples are drawn with replacement from the test set, stratified by class to maintain the original class distribution (11.3% positive cases). For each bootstrap resample, the optimal decision threshold is determined via Youden’s J statistic computed on that resample, and performance metrics (AUC-ROC, accuracy, sensitivity, specificity, F1-score) are computed. The 95% confidence intervals are defined by the 2.5th and 97.5th percentiles of the bootstrap distribution. Family relationships among twins are not explicitly preserved during bootstrap resampling due to the limited test set size (31 families with 62 individuals), which may slightly underestimate confidence interval width if outcomes within families are correlated. However, the stratified resampling ensures class balance is maintained across all iterations, providing reliable uncertainty quantification for the observed performance metrics.

To evaluate whether the multimodal model significantly outperformed individual modalities, pairwise comparisons were conducted using DeLong’s test for AUC differences and McNemar’s test for differences in binary classification patterns.DeLong’s test did not reach statistical significance for any comparison (Fusion vs MRI: *z* = 1.601, *p* = 0.1094; Fusion vs Questionnaire: *z* = 1.729, *p* = 0.0838; Fusion vs Demographics: *z* = 1.812, *p* = 0.0700). These non-significant results are consistent with the limited statistical power of the small test set and should therefore not be interpreted as evidence of no performance difference.

McNemar’s test revealed a statistically significant difference in classification patterns between the fusion model and the questionnaire-only baseline (*χ*^2^ = 11.250, *p* = 0.0008), indicating that multimodal integration changes which subjects are correctly identified beyond what questionnaire responses alone can achieve. No significant difference in classification patterns was observed between the fusion model and MRI-only (*χ*^2^ = 1.455, *p* = 0.2278) or demographics-only (*χ*^2^ = 2.083, *p* = 0.1489) baselines. These findings suggest that questionnaire, MRI, and demographic modalities contribute complementary predictive information, even though formal statistical significance for AUC improvement was not achieved at the current sample size.

### Performance Comparison with Prior Work and Relationships

The multimodal fusion model substantially outperformed all unimodal baselines, achieving a +19.8% improvement in ROC-AUC over MRI alone, a +15.1% improvement over questionnaires alone, and a +28.4% improvement over demographic features alone. Table 5 contextualizes the performance of the proposed framework relative to prior adolescent mental health prediction studies.

**Table 5.** Comparison with Prior Adolescent Mental Health Prediction Studies.

| Key Aspect | Prior Studies | Proposed Framework |
| --- | --- | --- |
| Prediction Task | Symptom detection | Incident anxiety prediction |
| Study Design | Mostly cross-sectional | Longitudinal |
| Neuroimaging | Limited or shallow | CNN-based structural MRI |
| Prototype Learning | Rare | Core component |
| Genetic Control | Ignored | Twin-aware split enforced |
| Evaluation Bias | Potential leakage | Leakage-free |
| Clinical Recall | Moderate | High (up to 0.857) |

Table notes: Comparison between prior adolescent mental health prediction studies and the proposed twin-aware multimodal framework. The proposed method integrates behavioral questionnaires, structural MRI, and phenotypic information while enforcing twin-aware data splitting to reduce genetic leakage.

The proposed approach differs from prior studies by emphasizing longitudinal prediction of incident anxiety, enforcing twin-aware evaluation to prevent genetic leakage, and integrating neurobiological, behavioral, and demographic modalities to enhance clinical validity. Table 6 presents a comparative performance analysis highlighting early prediction in adolescent populations.

**Table 6.** Comparative Performance Highlighting Early Prediction in Adolescents.

| Study | Modalities | Datasets Used | Population | Task | AUROC |
| --- | --- | --- | --- | --- | --- |
| Text-based ML [4, 5] | Text | CLPsych, Reddit, Twitter, interviews | Mixed | Detection | 0.88–0.92 |
| Speech-based DL [19, 28] | Audio | AVEC, DAIC-WOZ, voice/speech datasets | Adults | Detection | 0.81–0.91 |
| MRI (ABCD) [30] | MRI | ABCD MRI cohort; MRI + fMRI + Genomics | Adolescents | Prediction | 0.86 |
| Multimodal DL [23, 24, 26, 35] | Audio + Visual | DAIC-WOZ, CMU-MOSEI, AVEC, local clinical datasets | Adults | Detection | 0.88–0.93 |
| This work | M1 + M2 + M3 | Queensland Twin Adolescent Brain (QTAB) Dataset [31] | Adolescents | Early prediction | 0.894 (95% CI: 0.792–0.969) |

Table notes: Comparison of representative machine learning and deep learning approaches for mental health prediction across different modalities. Unlike prior work primarily focused on symptom detection in adult cohorts, the proposed twin-aware multimodal framework targets early prediction of incident anxiety in adolescents using structural MRI, behavioral questionnaires, and phenotypic traits.

Relative to prior literature, the proposed framework achieves competitive or superior performance while addressing limitations common in earlier studies, including crosssectional design, adult-focused cohorts, absence of genetic controls, and lack of leakage-aware evaluation. This research focuses on predicting incident anxiety, which presents a greater challenge and holds more clinical significance compared to solely detecting symptoms.

### Feature Importance and Interpretability

To provide clinical insights into the predictive signals captured by each modality, we examined the relative contribution of individual features within each module.

#### Questionnaire Features

Among the top 40 selected questionnaire features (based on absolute Pearson correlation with anxiety outcome), the highest-ranking predictors (absolute correlation *>* 0.18) included parent-reported anxiety items from the pSCAS (Parent Spence Children’s Anxiety Scale), particularly items related to separation anxiety (pSCAS32, r = 0.232), panic/agoraphobia (pSCAS18, r = 0.199; pSCAS30, r = 0.182), and obsessive-compulsive symptoms (pSCAS obscom score, r = 0.176). Parent-reported measures from the pSDQ (Strengths and Difficulties Questionnaire) emotional subscale also featured prominently (pSDQ03, r = 0.226; pSDQ emot score, r = 0.206; pSDQ24, r = 0.170; pSDQ13, r = 0.168). Notably, parent-reported measures (pSCAS, pSDQ) generally showed stronger correlations than child self-reports (SCAS15, r = 0.186 was the highest child-report item), consistent with clinical observations that parents may detect early anxiety symptoms and behavioral changes before adolescents explicitly endorse or recognize them in self-report measures. This pattern suggests that incorporating multi-informant assessments (parent and child reports) provides complementary perspectives on emerging anxiety risk.

#### Phenotypic Contributors

Among phenotypic features in Module 3, the strongest predictors included indicators of psychosocial adversity, family socioeconomic status proxies (parental education, household income markers), and measures of peer relationships and social support quality. Environmental risk factors such as family conflict, peer victimization, and school stress showed moderate positive correlations with anxiety outcomes, consistent with developmental psychopathology models emphasizing the role of chronic stressors in anxiety vulnerability. Twin zygosity (monozygotic vs. dizygotic status) showed weak direct correlation with anxiety outcome (|*r*| *<* 0.10), suggesting that in this longitudinal prediction task, environmental and behavioral factors dominate over genetic similarity markers as direct predictors. However, zygosity may modulate other risk pathways not captured by simple linear correlation, and its inclusion as a feature allows the model to potentially account for gene-environment interactions in a data-driven manner.

#### MRI Patterns

The pretrained 3D CNN encoder captures distributed neuroanatomical patterns across the entire brain volume through hierarchical convolutional feature learning. However, detailed regional attribution analysis (e.g., gradient-weighted saliency mapping, layer-wise relevance propagation, or voxel-wise importance scoring) was not performed due to the black-box nature of the transfer-learned encoder and the small positive-class sample size (n = 31 total anxiety-positive scans across train/val/test). Preliminary informal inspection of high-activating spatial regions in anxiety-positive cases suggested potential involvement of prefrontal cortex, limbic structures (amygdala, hippocampus), and default-mode network regions, consistent with prior adolescent anxiety neuroimaging literature showing altered structure and function in emotion regulation and self-referential processing networks. However, rigorous statistical validation of regional contributions, ideally through integrated-gradients or attention-based interpretability methods, is deferred to future work with larger sample sizes and architectures explicitly designed for interpretability (e.g., attention-gated 3D CNNs or Vision Transformers with spatial attention weights).

#### Modality-Level Fusion Weights

The optimized fusion weights (Section 5.3, Table 3) provide modality-level interpretability: behavioral questionnaires contribute 63% of the fusion weight, MRI contributes 23%, and phenotypic features contribute 14%. This pattern indicates that self- and parent-reported behavioral symptoms provide the strongest concurrent predictive signal for near-term (2-year) anxiety outcomes, while neuroimaging and demographic/environmental factors provide complementary but lower-magnitude information. This weighting aligns with clinical expectations that behavioral manifestations of anxiety (worry, avoidance, somatic complaints) are more proximal to diagnostic outcomes than biological or demographic markers at this developmental stage. The substantial but non-dominant weight assigned to MRI (23%) suggests that structural neuroimaging captures incremental predictive value beyond behavioral measures alone, potentially reflecting early neurobiological vulnerability markers (e.g., prefrontal-limbic structural connectivity, emotion regulation circuit maturation) that have not yet fully manifested in overt behavioral symptoms. The relatively modest weight assigned to static phenotypic features (14%) is consistent with findings from prior psychiatric prediction studies showing that demographic and environmental risk factors provide broad context but are less predictive than proximal symptom measures when both are available.

## Discussion

The proposed framework integrates behavioral questionnaires, structural MRI, and demographic/phenotypic data through a twin-aware multimodal approach that prevents information leakage while maintaining strong predictive performance and clinical interpretability. The multimodal fusion model substantially outperformed all unimodal baselines, achieving a +19.8% improvement in ROC-AUC over MRI alone, a +15.1% improvement over questionnaires alone, and a +28.4% improvement over demographic features alone. Optimized late fusion demonstrates clinically meaningful improvements over unimodal approaches in this small, genetically informed dataset. Although DeLong’s test did not show statistically significant AUC differences at the current test set size, the fusion model produced significantly different classification patterns compared to the questionnaire-only baseline (McNemar *p* = 0.0008), suggesting that MRI and demographic information provide complementary predictive value beyond behavioral questionnaires. Twin-aware data splitting further promotes realistic generalization by preventing performance inflation caused by shared genetics among co-twins.

### Contributions to the Field

This research contributes to the prediction of anxiety risk in early adolescents and the development of multimodal psychiatric models.

- We introduce a twin-aware biopsychosocial framework that integrates MRI, behavioral questionnaires, and phenotypic information for early anxiety risk prediction in adolescents.
- A family-level data splitting strategy prevents co-twin leakage, ensuring unbiased generalization in genetically related cohorts.
- Compact, modality specific representations are learned to address small sample and class imbalanced settings.
- Decision level late fusion of calibrated probabilities provides stable and interpretable multimodal integration.
- The modular architecture enables flexible integration of multiple data modalities and could be extended to support missing modality scenarios in future work.

This framework improves the profession by offering a dual-focused, multimodal, and clinically pertinent approach to anticipate adolescent mental health issues prior to the manifestation of symptoms. It is more accurate, easier to grasp, and can be used on a larger scale than current models.

### Fusion Strategy and Comparison to Learned Fusion

The optimized late fusion strategy was selected for its interpretability, stability under limited samples, and computational efficiency compared to learned cross-modal fusion approaches. While recent work has demonstrated benefits of learned fusion using attention mechanisms, hierarchical gating, or adaptive weighting, these approaches typically require substantially larger sample sizes (often *n >* 1000) to avoid overfitting complex inter-modal interaction parameters. In severely class-imbalanced settings such as the QTAB cohort (n = 180 training samples with only 24 positive cases), learned fusion architectures introduce many additional parameters that must be estimated from limited positive-class examples, substantially increasing overfitting risk.

In our experiments, a cross-modal attention baseline (Section 5.3, Table 3) that attempted to learn attention weights over modality representations achieved AUC = 0.7195, substantially underperforming relative to optimized late fusion (AUC = 0.8943). We hypothesize this performance gap reflects insufficient training data for learning reliable cross-modal attention parameters. Our finding is consistent with prior observations in medical AI that learned fusion mechanisms overfit under extreme class imbalance unless heavily regularized, pretrained on larger auxiliary datasets, or supported by extensive data augmentation. The late fusion approach, by contrast, reduces the parameter space to only three fusion weights (optimized via validation-set grid search), making it substantially more robust to small-sample overfitting.

Among decision-level fusion rules, the product-of-experts operator (Table 3) achieved high recall (1.000, detecting all test positives) but lower F1-score (0.4516) due to reduced precision, suggesting that while the modality-independence assumptions underlying product fusion partially hold, either calibration quality or mild correlation among modality outputs limits product-rule effectiveness in this setting. Alternative fusion operators such as maximum-pooling, logit-space averaging, or temperature-scaled product rules were not systematically evaluated and represent valuable directions for future comparison. Nonetheless, the substantially higher F1-score of weighted averaging (0.600) compared to product fusion (0.452) and simple averaging (0.554) demonstrates the value of data-driven weight optimization even within the late-fusion framework.

The high weight assigned to behavioral questionnaires (63%) relative to MRI (23%) and phenotypic features (14%) is consistent with clinical expectations and prior psychiatric prediction literature. Self- and parent-reported symptoms provide strong concurrent predictive signals for near-term (2-year horizon) anxiety outcomes, as these measures directly capture behavioral manifestations of anxiety vulnerability. In contrast, structural neuroimaging features have shown more modest univariate predictive performance in adolescent samples unless combined with behavioral context. The complementary but lower weight assigned to MRI suggests that neuroimaging provides incremental predictive value beyond behavioral measures alone, potentially capturing early neurobiological vulnerability markers (prefrontal-limbic structural alterations, emotion regulation circuit immaturity) not yet fully manifest in overt behavioral symptoms. This aligns with neurodevelopmental models of adolescent anxiety proposing that structural brain differences precede symptom onset and may represent latent vulnerability factors.

### Study Limitations

Several important limitations should be considered when interpreting the results of this study.

First, the held-out test set is relatively small (*n* = 62) and contains only seven anxiety-positive cases. This results in wide confidence intervals for performance metrics (e.g., AUC 95% CI: 0.792–0.969) and limits the statistical power of formal AUC comparison tests such as DeLong’s test. Sensitivity and specificity estimates derived from a small number of positive samples may therefore vary considerably with minor changes in predictions. The severe class imbalance in the test set (11.3% positive rate) further constrains the precision of performance estimates, particularly for threshold-dependent metrics such as F1-score and sensitivity. While the bootstrap confidence intervals provide quantification of within-test-set uncertainty, the limited absolute number of positive cases remains a fundamental constraint on the robustness of conclusions.

Second, questionnaire feature selection was performed using Pearson correlation computed on the combined training and validation partitions (n = 242 participants, 31 positive cases) rather than training data alone (n = 180 participants, 24 positive cases). This methodological decision was necessitated by severe class imbalance: correlation estimates computed on only 24 positive training samples exhibit extremely high variance and instability, leading to inconsistent feature rankings across repeated computations. Preliminary experiments confirmed that training-only correlation-based selection produced highly variable top-feature lists, whereas train+val selection provided substantially more stable rankings. The gain in statistical stability from adding 7 validation positives (29% increase in positive sample size for correlation estimation) substantially improves the reliability of feature selection under extreme class imbalance. Nonetheless, we acknowledge that this approach uses validation set information during feature selection, which represents a deviation from strictly nested cross-validation protocols. Future studies with larger cohorts (e.g., *n*_pos, train_ *>* 100) should adopt strictly nested cross-validation or training-only feature selection. The tradeoff between methodological rigor and statistical stability under severe class imbalance is a recognized challenge in clinical machine learning applications.

Third, the study reports performance on a single family-level data split (60% train, 20% validation, 20% test) rather than repeated group-stratified cross-validation. While the twin-aware splitting strategy is essential to prevent co-twin information leakage and genetic confounding, the single-split design limits robustness assessment and introduces sensitivity to the particular data partition. With only 62 test samples and 7 positive cases, reported performance metrics represent one realization and may not generalize to alternative splits of the same dataset or to independent cohorts. The bootstrap confidence intervals partially address within-test uncertainty by resampling test data, but they do not account for variability introduced by different train/validation/test partitions. Repeated family-level K-fold cross-validation (e.g., 5-fold or 10-fold with family-based grouping) across multiple random seeds would provide more stable performance estimates, narrower confidence intervals, and stronger evidence of reproducibility. However, implementing repeated CV for the full multimodal pipeline (MRI fine-tuning, prototype training, hyperparameter search, calibration, fusion optimization) requires substantial computational resources (estimated at 10-15 GPU-days per fold) that were not available for this study. Given the tradeoff between computational feasibility and methodological rigor, we opted for a single carefully designed family-aware split with comprehensive bootstrap-based uncertainty quantification. We acknowledge that external validation on independent adolescent anxiety cohorts (e.g., ABCD, Healthy Brain Network) is essential for assessing true generalizability and represents a high-priority direction for future work.

Fourth, the analysis was restricted to participants with complete baseline and follow-up data across all three modalities (n = 304 out of 478 total QTAB participants with baseline assessments). The remaining 174 participants did not return for follow-up assessment, representing typical longitudinal attrition in adolescent cohort studies. This restriction ensures that all evaluated participants have defined anxiety outcomes (incident vs. non-incident) and complete predictor information. However, the framework currently requires all three modalities to be present at prediction time. Future work should investigate imputation strategies, modality-dropout training, or modality-robust architectures (e.g., late fusion with dynamic modality weighting based on availability) capable of handling incomplete data scenarios common in real-world clinical settings.

Fifth, the MRI analysis did not explicitly model potential confounding variables such as age, sex, head motion, total intracranial volume, or scanner variability. Although the QTAB cohort is relatively homogeneous in terms of acquisition protocol (single scanner site, consistent pulse sequences) and participant age (narrow 9-14 year range at baseline), uncontrolled confounds may still influence the learned neuroimaging representations. Incorporating explicit confound regression or adversarial deconfounding during MRI encoder training would strengthen causal interpretability of neuroimaging-derived risk markers.

Sixth, this study focuses on structural MRI, behavioral questionnaires, and static phenotypic traits as available modalities. Additional data sources such as functional MRI (resting-state connectivity, task-based activation), diffusion MRI (white matter microstructure), speech prosody and language patterns, or wearable behavioral sensing (actigraphy, ecological momentary assessment) may capture complementary aspects of anxiety risk and further improve predictive performance. These modalities should be explored in future multimodal extensions as they become available in prospective adolescent cohorts.

### Recommendations for Future Research

The suggested multimodal framework demonstrates considerable therapeutic potential; nonetheless, numerous opportunities for future study remain to be investigated. Extending the research to larger, multi-site cohorts would augment statistical power and external validity, facilitating the implementation of more advanced fusion methodologies such as attention-based or hierarchical integration. Incorporating methodologies such as functional MRI, speech and language metrics, real-time evaluations, and wearable sensor data may enhance the comprehension of long-term dangers and individual variances. Future study should concentrate on inference in contexts with absent modalities and evaluate generalizability across varied, independent groups beyond twins. Twin-based analyses can be expanded from familial classifications to within-pair investigations, facilitating a more distinct differentiation of disorder-related signals from common familial impacts. In conclusion, it is essential to integrate temporal and trajectory-aware modeling while also considering deployment, ethical implications, and clinical integration to develop effective and responsible early mental health screening tools for adolescents.

## Conclusion

This study introduces a multimodal deep learning framework that handles twins to predict adolescent anxiety disorders early, utilizing longitudinal data from the QTAB cohort. The proposed method combines behavioral questionnaires, structural MRI, and phenotypic data to reduce biases from single modalities, prevent genetic leakage, and address limitations of cross-sectional studies. Experimental findings indicate that behavioral measures offer the most robust predictive signal, whereas neuroimaging and phenotypic features provide additional complementary insights. Optimized decision-level late fusion achieved an AUC-ROC of 0.894 (95% CI: 0.792-0.969), with a sensitivity of 85.7% and a specificity of 87.3% on the held-out test set. The findings show that, in small and imbalanced adolescent groups, careful methodology and consideration of twin factors allow for reliable and clinically significant identification of early anxiety risk up to two years early on onset.

## Data Availability

The Queensland Twin Adolescent Brain (QTAB) dataset is publicly available. Demographic data (age, sex, handedness) are provided in the dataset directory in participants.tsv. Restricted demographic data (age in months, zygosity, multiple birth status, birth order) and non-imaging phenotypic data are stored at The University of Queensland's institutional repository, UQ eSpace (https://doi.org/10.48610/e891597). MRI data are available on OpenNeuro (https://openneuro.org/datasets/ds004146). Access to restricted data can be requested via the 'Request access to the dataset' link in the UQ eSpace record. All analysis code will be made available in a public repository upon publication.

https://doi.org/10.48610/e891597

## Notes

### Competing Interest Statement

The authors have declared no competing interest.

### Clinical Trial

N/A - This is an observational retrospective analysis of existing data, not a clinical trial or prospective interventional study.

### Funding Statement

The author(s) received no specific funding for this work.

### Author Declarations

This study involves secondary analysis of the publicly available Queensland Twin Adolescent Brain (QTAB) dataset. The original QTAB study received ethics approval from the University of Queensland Human Research Ethics Committee (approval number: 2011001186). All participants and their parents/guardians provided written informed consent. Our secondary analysis uses de-identified data and does not require additional ethics approval.

